# An Observational Study of COVID-19 from A Large Healthcare System in Northern New Jersey: Diagnosis, Clinical Characteristics, and Outcomes

**DOI:** 10.1101/2020.08.07.20170357

**Authors:** Yanan Zhao, Marcus H. Cunningham, Jose R. Mediavilla, Steven Park, Sean Fitzgerald, Hee Sang Ahn, Xiangyang Li, Caixin Zhan, Tao Hong, Gary Munk, Kar Fai Chow, David S. Perlin

**Affiliations:** Center for Discovery and Innovation, Hackensack Meridian Health, Nutley, NJ, USA; Department of Medical Sciences, Hackensack Meridian School of Medicine, Nutley, NJ, USA; Molecular Diagnostic Laboratory, Department of Pathology, Hackensack University of Medical Center, Hackensack, NJ, USA; Microbiology Laboratory, Department of Pathology, Hackensack University of Medical Center, Hackensack, NJ, USA; Department of Pathology, Hackensack Meridian School of Medicine at Seton Hall University, Nutley, NJ, USA; Core Laboratory, Department of Pathology, Hackensack University of Medical Center, Hackensack, NJ, USA; Clinical Virology Laboratory, Department of Pathology, Hackensack University of Medical Center, Hackensack, NJ, USA

**Keywords:** COVID-19, clinical characteristics, outcome, Ct value, viral load

## Abstract

**Background:** New Jersey was an early epicenter for the COVID-19 pandemic in the United States, yet information on hospitalized COVID-19 patients from this area is scarce. This study aimed to provide data on demographics and clinical features of a hospitalized patient population who were confirmed with infection by our in-house (CDI) real-time reverse-transcription polymerase chain reaction (RT-PCR) test.

**Methods:** We included consecutive patients who were admitted to Hackensack Meridian Health system hospitals with laboratory-confirmed diagnoses of COVID-19 at Hackensack University Medical Center by the CDI virus test between March 12, 2020, and April 8, 2020. Clinical data and viral testing results were collected and analyzed for characteristics associated with outcomes, as well as the correlation with viral load.

**Results:** A total of 722 patients were included in the study, with a median age of 63 (interquartile range (IQR), 51-75) and 272 (37.7%) females. Mortality of this case series was 25.8%, with a statistically significant linear increase observed from age 40 to ≥80 by 10-year intervals. Viral load, as indicated by the cycle of threshold (Ct) values from the RT-PCR test, was significantly higher in the oldest patient group (≥80), and inversely correlated with survival.

**Conclusions:** This is the first report to describe the clinical characteristics and outcomes in a large hospitalized COVID-19 patient series from New Jersey. Findings from this study are valuable to the ongoing response of both nationwide healthcare networks and the medical research community.

**Summary:** We describe the diagnosis, clinical characteristics, and outcomes of a large hospitalized patient population in northern New Jersey during the early stages of the COVID-19 pandemic.

## INTRODUCTION

The 2019 novel coronavirus disease (COVID-19) caused by severe acute respiratory syndrome coronavirus 2 (SARS-CoV-2) was first identified in Wuhan, China in December 2019 [1]. Since then it has quickly spread globally and was declared a pandemic by the World Health Organization (WHO) in March 2020 [2, 3]. The first confirmed case of COVID-19 in the United States was reported on January 20, 2020 [4]. Thereafter, the virus spread quickly throughout the US, as New York and New Jersey became the earliest epicenters of the disease.

As the largest private health system in NJ, Hackensack Meridian Health (HMH) responded quickly and professionally to this unprecedented public health crisis. During the early stages of the pandemic, HMH’s response was challenged by the initial limitations of the SARS-CoV-2 reverse-transcription polymerase chain reaction (RT-PCR) test developed by the US Centers for Disease Control and Prevention (CDC) (https://www.cdc.gov/coronavirus/2019-ncov/about/testing.html, accessed on July 23, 2020) and extensive delays (7-10 days) in test results, highlighting the urgent need to establish an accurate and practical diagnostic method in local hospitals in the period of time when no commercial test was approved or available. We systematically evaluated both CDC diagnostic panel and another real-time RT-PCR diagnostic panel developed by researchers in Germany [5]. The latter had already adopted by the WHO (referred throughout as WHO panel) as their official molecular diagnostic panel for COVID-19 (https://www.who.int/emergencies/diseases/novel-coronavirus-2019/technical-guidance/laboratory-guidance), and has been widely used in many European laboratories [6]. Based upon our evaluation results, we built a hybrid diagnostic panel (CDI Enhanced COVID-19 Test) and quickly implemented the test in the molecular laboratory of Hackensack University Medical Center (HUMC), upon acquisition of the pre-Emergency Use Authorization (EUA) approval from the US Food and Drug Administration (FDA) on March 12, 2020. Until April 9, the CDI test was the only test used at HUMC to diagnose COVID-19, after which multiple commercial diagnostic platforms (ID NOW™ COVID-19 test, cobas® SARS-Cov-2 test, Xpert® Xpress SARS-CoV-2, BioFire COVID-19 test) became available in the laboratory to expand testing capacity.

As the pandemic continues to unfold, data involving the clinical characteristics and outcomes of COVID-19 disease are emerging [7-12]. However, information regarding infections early in the NJ outbreak is scarce. In this study, we describe for the first time the diagnostic, demographic and clinical characteristics of COVID-19 patients hospitalized within the HMH system and confirmed the infection by the CDI test at HUMC at the earliest stage of the epidemic in New Jersey.

## METHODS

### Patient population

In this retrospective, observational study, we included consecutive patients admitted to HMH system hospitals with laboratory confirmed COVID-19 diagnoses at the HUMC Molecular Diagnostic Laboratory utilizing the CDI-enhanced RT-PCR test between March 12, 2020, and April 8, 2020. Clinical outcomes were monitored until May 22, 2020, the final date of follow-up. The study was approved by the HMH institutional review board.

### Data collection

Clinical data including demographic background, comorbidities, manifestation, therapeutic options, and clinical outcomes were retrieved from the real-world database built through the HMH health record system and provided to the study team in a de-identified fashion.

### Laboratory confirmation of COVID-19

The CDI-enhanced COVID-19 test was utilized to detect the presence of SARS-CoV-2 RNA in nasal and/or throat swab specimens collected from patients. The test was approved for use on March 12, 2020 under FDA Emergency Use Authorization for COVID-19 (https://www.fda.gov/media/137036/download). This test panel includes two virus detection components, the E and N2 assays, targeting the envelope and nucleocapsid protein genes of SARS-CoV-2, respectively. The limit of detection was less than 20 viral genome copies per reaction. The third component included in the diagnostic panel is the RP assay, which targets the human RNase-P gene as an internal control for sample quality evaluation. The test was performed at the Molecular Diagnostic Laboratory of HUMC, following the standard operating procedure as published on the FDA website. Briefly, 200 μl of nasopharyngeal or oropharyngeal swab sample was used for total nucleic acid (TNA) extraction by the MagNA Pure 24 system (Roche Life Science) according to manufactural instructions. TNA samples were immediately subjected to the RT-PCR test. The primer/probe sequences and the RT-PCR setup protocols are detailed in the **supplementary material**. The cycle of threshold (Ct) value was noted at the end of each test and recorded for all reactions included in the run. The Ct value cutoff for sample positivity was 40 cycles.

### Statistical analysis

Statistical analyses were carried out using SPSS version 17.0, and graphs were plotted using GraphPad Prism version 8.4.2. Continuous and categorical variables were presented as median (interquartile range [IQR]) and n (%), respectively. We used the Mann-Whitney U test, χ^2^ test, or Fisher’s exact test to compare differences between survivors and non-survivors where appropriate. Phi, Pearson, or Spearman’s correlation was used to assess the relationships between different variables as appropriate. A p value less than 0.05 was considered statistically significant.

## RESULTS

A total of 722 patients were included in the study, with a median age of 63 (IQR, 51-75), of which 272 (37.7%) were females (**Table 1**). Of these, 716 were adult patients (age ≥18), while the rest included 3 infants (age <1) and 3 pediatric patients. Among all racial/ethnic categories, white (363, 50.7%) was the most prevalent, followed by Hispanic (143, 20.0%). Healthcare workers accounted for 12.7% (92/722) of this case series. The most common morbidities were hypertension (373, 52.2%), obesity (249, 40.4%) and diabetes (210, 29.5%). Fever (520, 73.0%) was the most common symptoms at admission, followed by shortness of breath (507, 70.9%) and cough (487, 68.4%). All patients were tested positive for SARS-CoV-2 by the CDI-enhanced COVID-19 test. The median time from test order (sample collection) to report was 19.7 hours (IQR, 9.1-26.9). The vast majority of patients (719, 99.6%) were positive on the initial diagnostic test, and only 3 patients had a negative initial test followed by a positive repeat test. The average time from admission to diagnosis was 21.8 hours.

**Table 1.** Demographics and Baseline Characteristics of Patients admitted to HMH hospitals and laboratory confirmed with COVID-19 by CDI enhanced COVID-19 test.

|  | No. (%) |
| --- | --- |
| Demographics |  |
| Total No. | 722 |
| Age, median (IQR) [range], years | 63 (51-75) [0-101] |
| Sex |  |
| Female | 272 (37.7) |
| Male | 450 (62.3) |
| Race/Ethnicity |  |
| No. | 716 |
| African American | 62 (8.6) |
| Asian | 36 (5.0) |
| White | 363 (50.7) |
| Hispanic | 143 (20.0) |
| Other | 112 (15.6) |
| Healthcare worker | 92 (12.7) |
| Former/current smoker [total No.] | 173 (25.5) [679] |
| Comorbidities [total No.] |  |
| Cancer | 112 (15.8) [707] |
| Diabetes | 210 (29.5) [711] |
| Asthma | 79 (11.2) [705] |
| COPD | 49 (7.0) [705] |
| Renal failure | 46 (6.6) [701] |
| HIV | 2 (0.3) [722] |
| Hepatitis | 11 (1.5) [722] |
| Hypertension | 373 (52.2) [714] |
| Heart failure | 52 (7.4) [703] |
| Adult BMI ≥ 30 | 249 (40.4) [617] |
| Presenting symptoms [total No.] |  |
| Fever | 520 (73.0) [712] |
| Shortness of breath | 507 (70.9) [715] |
| Cough | 487 (68.4) [712] |
| Nausea | 82 (11.4) [722] |
| Diarrhea | 121 (16.8) [722] |
| Loss of taste or smell | 11 (1.5) [722] |

IQR = interquartile range; COPD = chronic obstructive pulmonary disease; BMI = body mass index

The number of deaths in this case series was 186, registering an overall of 25.8% mortality rate. Yet, mortality was not evenly distributed among different age groups (**Fig. 1**). There was no recorded mortality in patients younger than 18 years old. Among adult patients, limited mortality was noted within the 18-29 (2 deaths) and 30-39 (0 deaths) age groups. By contrast, there was a statistically significant trend (linearity R^2^=0.9242, p=0.0091) of mortality gradually increasing from 6.5%, 12.9%, 20.7%, 38.0%, to 62.5% when the age group increased from 40 to ≥80 by 10-year intervals. Mortality was not significantly different between males (26.7%) and females (24.3%) (p=0.484). During hospitalization, 481 (66.6%) patients received treatment with hydroxychloroquine (**Table 2**). Antibiotics were given to 463 (64.1%) patients, with azithromycin being the most common. Fewer patients received treatments with corticosteroids (6.0%), IL-6 inhibitor (tocilizumab (2.9%) or sarilumab (0.4%)), or remdesivir (2.4%). There were 262 (36.3%) patients who required oxygenation support, while the percentage of oxygenation device use was significantly higher in non-survivors than that in survivors (44.6% vs. 33.4%, p=0.0078). Similarly, ventilator usage was much higher in non-survivors compared to survivors (45.7% vs. 7.3%, p<0.0001). Intensive care unit (ICU) admission rate was also substantially higher in non-survivors compared to survivors (57.5% vs. 16.0%, p<0.0001), although the duration of ICU stay was not significantly associated with death. The median hospital length of stay was 7 days (IQR, 4-13), with 11 days (IQR, 6-17) for non-survivors and 6 days (IQR, 3-11) for survivors.

**Figure 1.**
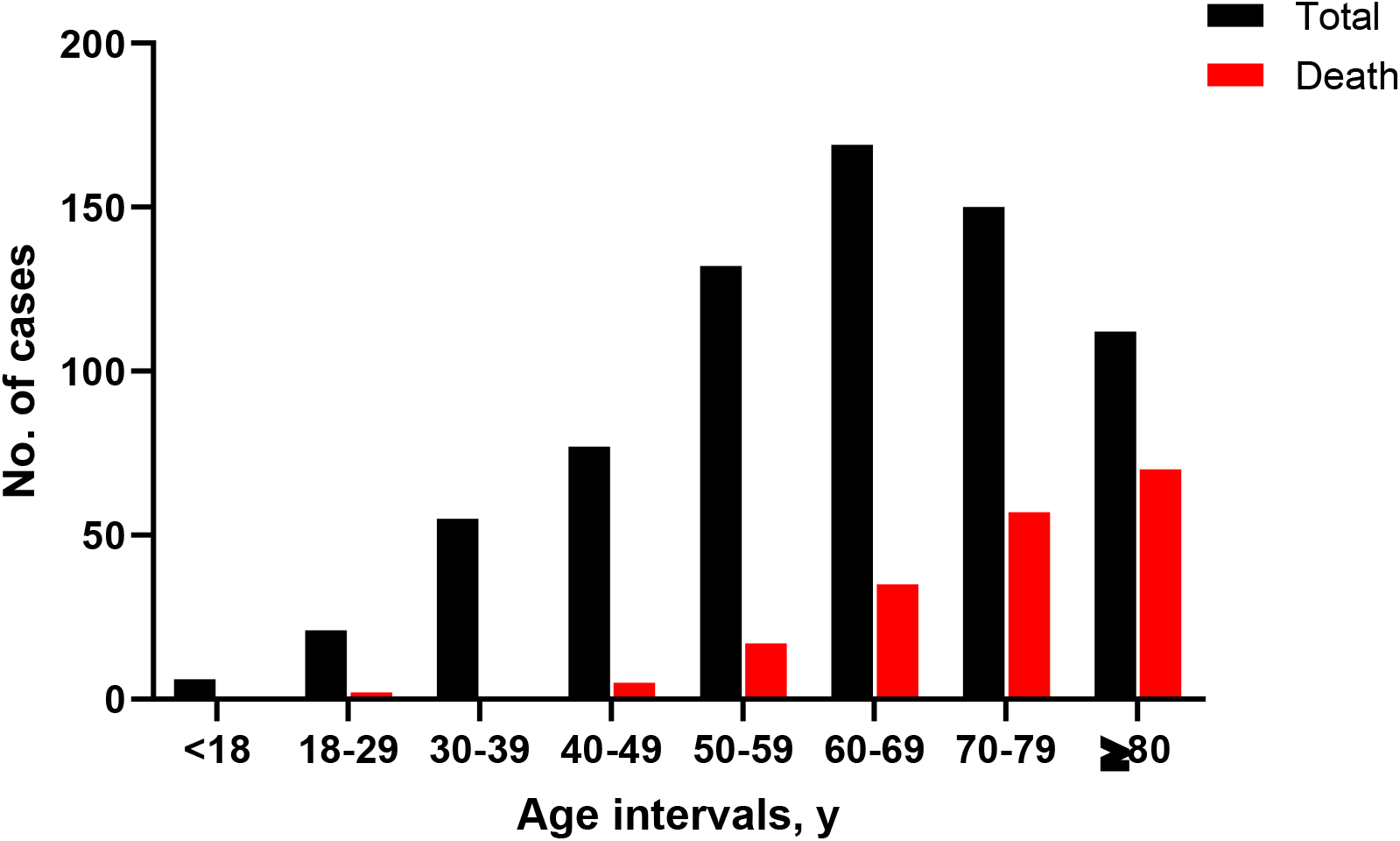
Case and death distribution among age groups.

**Table 2.**
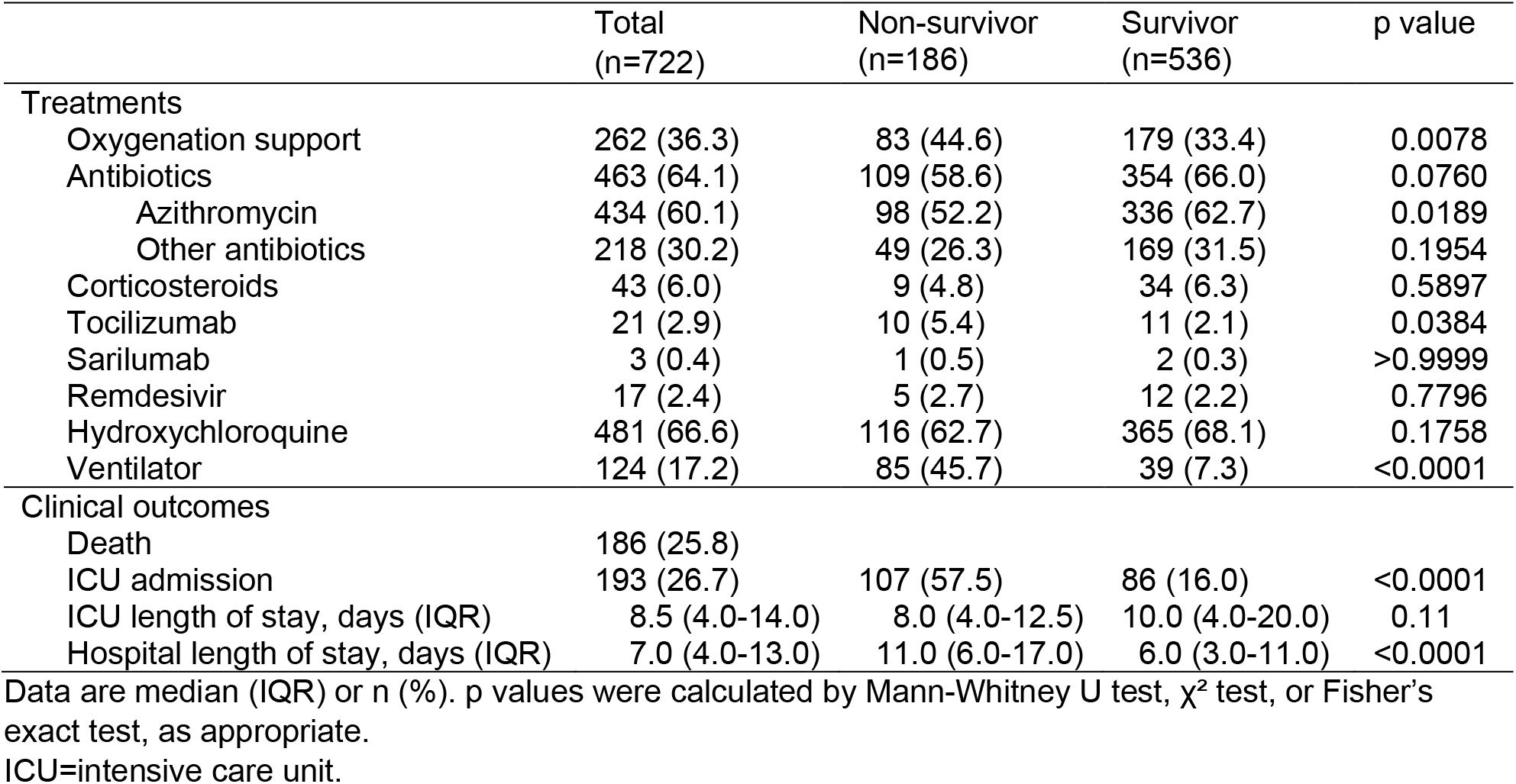
Treatment received during hospitalization and clinical outcomes.

Given the quantitative nature of the RT-PCR assay and the standardized procedure of our testing protocol, we interpreted the Ct values of E and N2 assays as a measure of viral load and investigated their correlation with patient demographics and clinical outcomes. The scatter plots (**Fig. 2**) of all Ct values and their distribution by different age groups for each assay demonstrated highly similar distribution patterns for both viral detection targets. The median Ct values for patients younger than 18 were 22.84 (IQR, 18.49-30.30) for the E assay and 23.43 (IQR, 18.53-34.48) for the N2 assay. Due to the very small sample size (n=6), patients younger than 18 were excluded from the Ct value comparison between different age groups. Among adults, patients ≥80 in age had median Ct values of 23.12 (IQR, 18.51-27.29) for the E assay and 23.78 (IQR, 19.54-29.18) for the N2 assay, significantly lower than those of any other adult group, and suggestive of higher viral load (p values listed on the table of Fig. 2). The average median Ct value difference between the oldest age group and all others was 4.22 for E and 5.04 for N2, indicating that the respiratory viral load carried by patients ≥80 was roughly 21~58-fold higher than that captured by other adult age group, based on the log-linear relationship between Ct value and viral RNA copy number established for each assay during the CDI assay development (data not shown).

**Figure 2.**
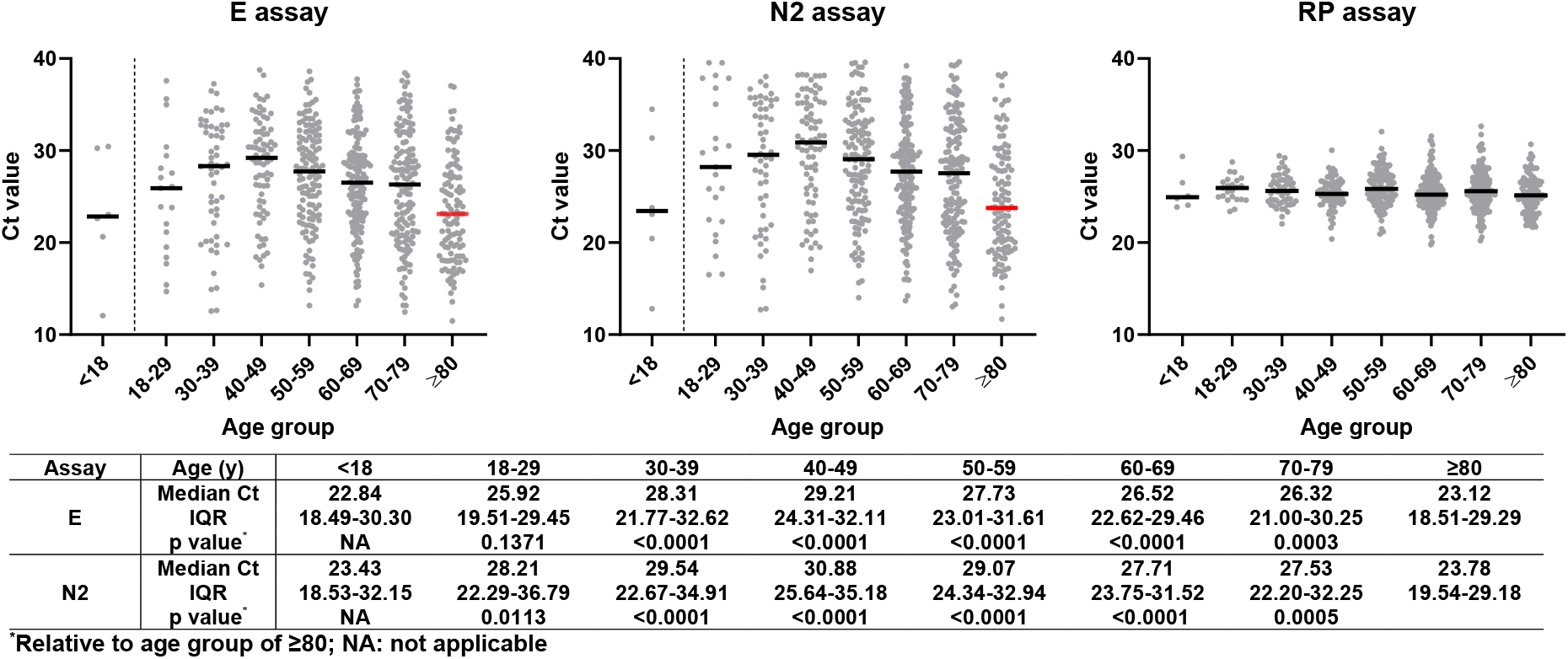
Scatter plots of Ct value distributions by age groups for E, N2, and RP assay. Solid horizontal short lines represent median Ct values for designated age groups. Patients ≥80 years old had lowest median Ct values for both virus detection assays (E and N2), compared to other adult age groups (table underneath the plots). Ct values from the quality control assay (RP) were very similar among different groups, suggesting overall good laboratory practice from sample processing to RT-PCR set up.

In order to understand the relationship between viral load and clinical outcome, we plotted the Ct values by patient survival (**Fig. 3**). The clear difference in Ct values between survivors and non-survivors demonstrated a striking association between viral load and survival (Spearman’s ρ= −0.271 for E assay Ct and death, p <0.001; Spearman’s ρ= −0.252 for N2 assay Ct and death, p <0.001). The median Ct values in non-survivors were lower by 4.67 (23.02 vs. 27.69) for E and 4.88 (24.03 vs. 28.91) for N2 relative to survivors, suggesting that the viral loads at diagnosis in non-survivors were approximately 29~50-fold higher. Notably, there were 6 patients in this case series who received at least one additional viral test during hospitalization following diagnosis. We followed up the Ct value dynamics and survival of these patients as shown in Fig. 4. One patient (#C) died 11 days after admission, while all other patients survived. A visible downward trend of viral load was observed in 4 out of 5 surviving patients, with concomitant Ct value increases (both E and N2) from an average of 24 cycles at diagnosis to ~35 or greater upon final testing. The only non-survivor had a fluctuating Ct profile over the 9-day course of follow-up. However, the fact that both E and N2 Ct values were very low at initial diagnosis, as well as upon final testing on day 9 post diagnosis demonstrated persistent infection by the virus.

**Figure 3.**
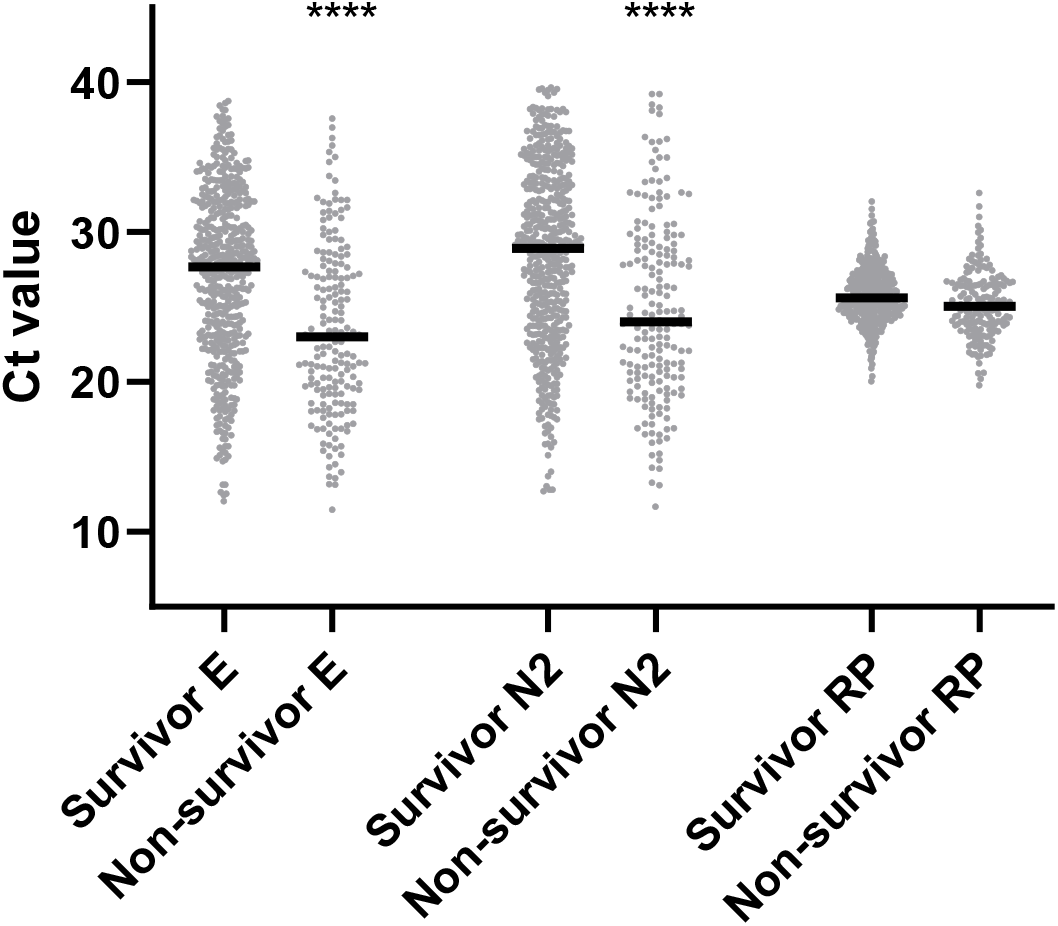
Ct value distribution by survival. Solid black lines represent median Ct values for designated age groups. Statistical significance for comparison between survivor and non-survivor was labeled by ****, p <0.0001.

**Figure 4.**
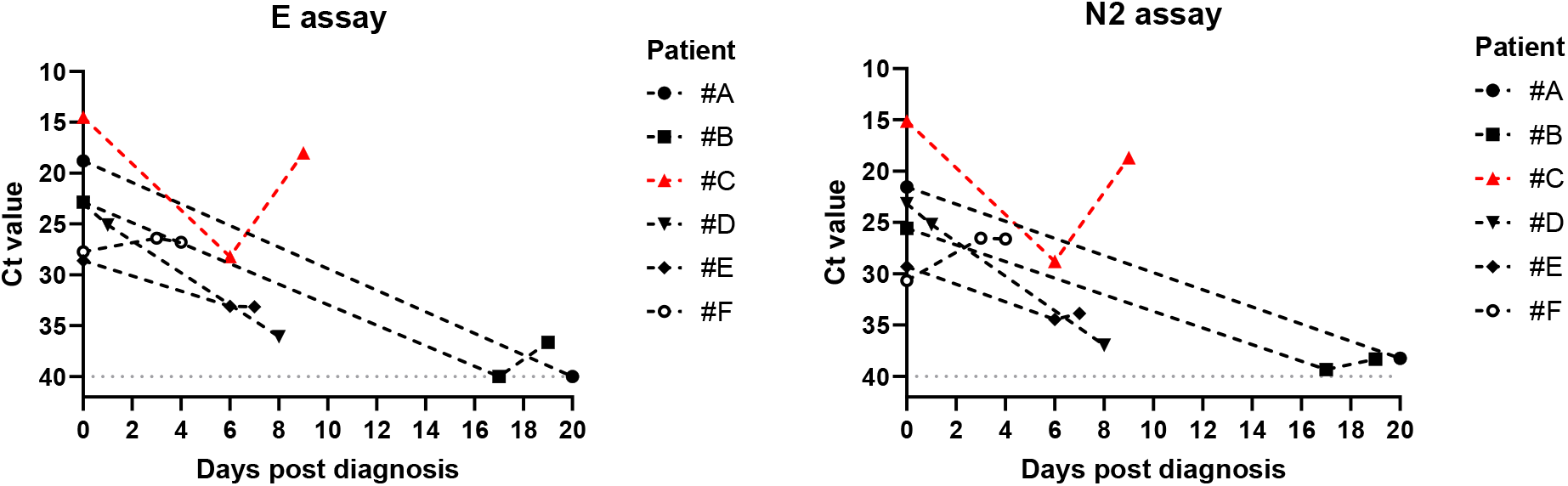
Dynamic changes in virus detection Ct values in patients who received multiple CDI COVID-19 tests after diagnosis during hospitalization. The x-axis uses a relative time scale, where each patient uses his/her own time of diagnosis as the starting point to record the time of each repeat test. Survived patients were shown in black markers and black lines, and non-survivor was shown in red. A virtual Ct value of 40 was used to plot negative detection results (patient #A on day 20 and patient #B on day 17) for E assay.

## DISCUSSION

To our knowledge, this is the first report to describe the clinical characteristics and outcomes in a large hospitalized patient series from NJ, during the early phase of the COVID-19 pandemic. The demographics of this case series were similar to those reported previously for patients hospitalized in the New York City area [13]. Healthcare workers accounted for more than 12% of the case series; however, conclusions cannot be made about the route or source of infection (nosocomial or community acquired) pending further investigation using different study designs. Pre-existing comorbidities were common, with more than half of the patients exhibiting hypertension prior to infection and over 40% obese. The mortality in our study was 25.8%, and a clear trend of progressive mortality increasing with age was apparent in patients ≥40 years old. Comparison of treatment received during hospitalization found that the most significant difference between survivors and non-survivors was the requirement for mechanical ventilation and oxygenation support. Only less than 1/3 of patients who needed ventilator support survived through the follow-up period. Other commonly used therapies including hydroxychloroquine, corticosteroids, and remdesivir were similarly distributed in survivors and non-survivors, except azithromycin and tocilizumab. The significance of a different proportion of tocilizumab usage was possibly due to the small number of patients enrolled in a clinical trial during the study period, therefore inconclusive. However, the use of azithromycin in a significantly lower proportion of non-survivors compared to survivors was somewhat convoluted, and further investigation on bacterial co-infections is warranted.

It is worth noting that this entire case series was confirmed to be COVID-19 positive by our in-house built, FDA approved, viral RT-PCR test. During the early phase (mid-March to early-April) of the pandemic in NJ, the availability of our in-house assay was extremely important, as the routine turnaround time for hospitals sending samples to the CDC or the state reference laboratory for COVID-19 test was around 5~7 days during a period of time when no commercial test kits were available. In the present study, most of the patients obtained a laboratory-confirmed COVID-19 diagnosis within 24 hours of admission, thereby enabling timely patient management and distribution of limited resources (e.g. negative pressure isolation rooms), while informing healthcare workers about potential risks for cross-infection.

Using Ct values acquired from the RT-PCR tests as an indicator of viral load in relation to clinical characteristics, we found that there was a considerably higher viral load in patients at age ≥80 at diagnosis than other adult patients, suggesting that the virus acquired by this age group replicates more actively in the host, and therefore may be more contagious. The viral load was also found to be significantly associated with death, and the average viral RNA copies detected in non-survivors were 29~50-fold higher than in survivors. Interestingly, a similar correlation was observed while tracing the Ct values from repeated viral tests in 6 individual patients, wherein viral load declined over treatment courses in 4 of 5 patients who survived. In contrast, the viral load in the only non-survivor remained high till the final test administered two days prior to death. These findings are consistent with previous reports [14-17], and further support the notion that viral load is an important factor associated with clinical outcome. Continuous high viral load may be a predictor of poor outcome.

Our study has several limitations. First, the study population only included patients who were confirmed to have SARS-CoV-2 infection by the CDI-enhanced test during the specified period of time. Due to the proximity to the laboratory performing the testing, patients from HUMC (n=604) accounted for 83.7% of the study population, while the rest were distributed in 11 other HMH hospitals. Second, clinical data were retrieved from a pre-designed database established for other purposes, and with missing values in multiple data fields. Third, given the retrospective observational nature of our study, no causal effect can be inferred, even if significant associations are observed.

In summary, we described the basic clinical features and outcomes of a patient population hospitalized within a large healthcare network during the early phase of the pandemic in NJ. In particular, we leveraged our experience developing an in-house viral testing platform towards patient management and infection control, which proved to be valuable and beneficial to both our healthcare network and the medical research community.

## Data Availability

NA

## ACKNOWLEDGMENTS

We thank Dr. Andrew Ip for providing the clinical information. We would also like to thank all medical staff who contributed to sample collection and transportation.

## REFERENCES

1. Zhu N, Zhang D, Wang W, et al. A Novel Coronavirus from Patients with Pneumonia in China, 2019. The New England journal of medicine 2020; 382(8): 727–33.

2. Coronavirus disease (COVID-19) pandemic. Available at: https://www.who.int/emergencies/diseases/novel-coronavirus-2019. Accessed march 25.

3. Coronavirus disease 2019 (COVID-19): situation summary. Available at: https://www.cdc.gov/coronavirus/2019-ncov/cases-updates/summary.html. Accessed March 25.

4. Holshue ML, DeBolt C, Lindquist S, et al. First Case of 2019 Novel Coronavirus in the United States. The New England journal of medicine 2020; 382(10): 929–36.

5. Corman VM, Landt O, Kaiser M, et al. Detection of 2019 novel coronavirus (2019-nCoV) by real-time RT-PCR. Euro surveillance: bulletin Europeen sur les maladies transmissibles = European communicable disease bulletin 2020; 25(3).

6. Reusken C, Broberg EK, Haagmans B, et al. Laboratory readiness and response for novel coronavirus (2019-nCoV) in expert laboratories in 30 EU/EEA countries, January 2020. Euro surveillance: bulletin Europeen sur les maladies transmissibles = European communicable disease bulletin 2020; 25(6).

7. Aggarwal S, Garcia-Telles N, Aggarwal G, Lavie C, Lippi G, Henry BM. Clinical features, laboratory characteristics, and outcomes of patients hospitalized with coronavirus disease 2019 (COVID-19): Early report from the United States. Diagnosis (Berlin, Germany) 2020; 7(2): 91–6.

8. Pereira MR, Mohan S, Cohen DJ, et al. COVID-19 in solid organ transplant recipients: Initial report from the US epicenter. American journal of transplantation: official journal of the American Society of Transplantation and the American Society of Transplant Surgeons 2020; 20(7): 1800–8.

9. Arentz M, Yim E, Klaff L, et al. Characteristics and Outcomes of 21 Critically Ill Patients With COVID-19 in Washington State. Jama 2020; 323(16): 1612–4.

10. Zhou F, Yu T, Du R, et al. Clinical course and risk factors for mortality of adult inpatients with COVID-19 in Wuhan, China: a retrospective cohort study. Lancet (London, England) 2020; 395(10229): 1054–62.

11. Chen N, Zhou M, Dong X, et al. Epidemiological and clinical characteristics of 99 cases of 2019 novel coronavirus pneumonia in Wuhan, China: a descriptive study. Lancet (London, England) 2020; 395(10223): 507–13.

12. Porcheddu R, Serra C, Kelvin D, Kelvin N, Rubino S. Similarity in Case Fatality Rates (CFR) of COVID-19/SARS-COV-2 in Italy and China. Journal of infection in developing countries 2020; 14(2): 125–8.

13. Richardson S, Hirsch JS, Narasimhan M, et al. Presenting Characteristics, Comorbidities, and Outcomes Among 5700 Patients Hospitalized With COVID-19 in the New York City Area. Jama 2020; 323(20): 2052–9.

14. He X, Lau EHY, Wu P, et al. Temporal dynamics in viral shedding and transmissibility of COVID-19. Nature medicine 2020; 26(5): 672–5.

15. Liu Y, Yan LM, Wan L, et al. Viral dynamics in mild and severe cases of COVID-19. The Lancet Infectious diseases 2020; 20(6): 656–7.

16. To KK, Tsang OT, Leung WS, et al. Temporal profiles of viral load in posterior oropharyngeal saliva samples and serum antibody responses during infection by SARS-CoV-2: an observational cohort study. The Lancet Infectious diseases 2020; 20(5): 565–74.

17. Zou L, Ruan F, Huang M, et al. SARS-CoV-2 Viral Load in Upper Respiratory Specimens of Infected Patients. The New England journal of medicine 2020; 382(12): 1177–9.

